# Delving into PubMed records: some terms in medical writing have drastically changed after the arrival of ChatGPT

**DOI:** 10.1101/2024.05.14.24307373

**Authors:** Kentaro Matsui

## Abstract

**Introduction:** It is estimated that large language models (LLMs) including ChatGPT is already widely used in academic paper writing. This study aims to investigate whether the usage of specific terminologies has increased, focusing on words and phrases frequently reported as overused by ChatGPT.

**Methods:** A list of 142 potentially AI-influenced terms was curated from online discussions and recent literature documenting LLM vocabulary patterns, while 84 common academic terms in the medical field were used as controls. PubMed records from 2000 to 2024 were analyzed to track the frequency of these terms. Usage trends were normalized using a modified Z-score transformation.

**Results:** Among the potentially AI-influenced terms, 100 displayed a meaningful increase (modified Z-score ≥ 3.5) in usage in 2024. The linear mixed-effects model showed a significant effect of potentially AI-influenced terms on usage frequency compared to common academic phrases (p < 0.001); the usage of potentially AI-influenced terms showed a noticeable increase starting in 2020.

**Discussion:** This study revealed that certain words, such as “delve,” “underscore,” “meticulous,” “boast,” and “commendable,” have been used more frequently in medical and biological fields since the introduction of ChatGPT. The usage of these terms had already been increasing prior to ChatGPT’s release, suggesting that ChatGPT accelerated the popularity of expressions already gaining traction. The identified terms can inform medical educators aiming to enhance awareness of language trends and promote best practices among trainees using LLMs.

## Introduction

ChatGPT rapidly achieved widespread global use after its launch on November 30, 2022. Trained on a vast corpus of text data, the large language model (LLM) including ChatGPT generates natural language with remarkable fluency. Shortly after its release, ChatGPT’s applicability for scientific writing in medical and biological fields became evident [1,2]. Due to the fervor surrounding its capabilities, it was credited as an author on several papers, igniting considerable debate (currently, AI is not acknowledged as an author in scholarly publications [3]). There were even opinions that the use of ChatGPT in paper writing was plagiarism [4], but in reality, LLMs such as ChatGPT, Gemini, and Claude are already being used in paper writing. The use of LLMs can be applied in various ways in academic writing [1,5] and is also important for the research activities of non-native researchers whose first language is not English [6-8]. Presently, a framework has been established that permits the use of LLMs in writing, provided their involvement is adequately acknowledged [3].

While LLMs can produce natural writing, their output also exhibits certain characteristics [9,10]. Recent reports focusing on detecting text generated by LLMs have identified several frequently used words, such as ‘commendable,’ ‘meticulous,’ ‘intricate,’ and ‘realm’ [11-14]. The extraction of these characteristic keywords of LLMs in these previous reports was performed by comparing human-generated text with ChatGPT-generated text [11,13,14].

While this approach revealed ChatGPT’s characteristics among the words commonly used by both humans and ChatGPT, it had methodological limitations in extracting words with low usage frequencies. Clarifying the word expressions that LLMs tend to use in medical and biological papers is crucial for designing academic writing support and medical education programs [15]. Moreover, revealing the extent of ChatGPT’s impact on papers in the medical and biological fields is essential for maintaining the fairness and reliability of academic research and from the perspective of research ethics [16]. However, the existing literature lacks a thorough investigation of the specific ways in which ChatGPT has transformed academic writing practices in the medical and biological disciplines, necessitating further research.

As the usefulness of LLMs becomes more evident, the number of researchers using LLMs for writing papers has been gradually increasing [11,13,17]. It would logically follow that there has been an increase in the number of research reports featuring specific expressions unique to LLMs. Despite anecdotal observations of LLMs favoring particular words like ‘delve,’ quantitative evidence regarding the increased usage of such specific, informally identified LLM-associated terms in scholarly publications remained scarce when this study was initiated. This study, therefore, tested the hypothesis that the adoption of certain scientific terminologies has risen following the advent of ChatGPT. Focusing on words and phrases frequently reported as overused by ChatGPT, I investigated PubMed records from 2000 onwards and performed a comparison using terms commonly used in medical research as a control. This analysis aims to empirically explore the influence of LLMs on the lexicon of medical literature.

## Materials and Methods

### Search for Records

To investigate the influence of LLMs on vocabulary usage in medical literature, I constructed a comprehensive list of potentially AI-influenced terms using a two-step approach. First, as an exploratory seed step, potentially AI-influenced terms were manually curated from publicly accessible online discussions (e.g., social media platforms, blogs, forums). Terms were included if independently identified by at least two different users as ‘overused by ChatGPT or other LLMs,’ across multiple online sources. Terms that appeared to be proper nouns, such as disease names, were excluded. Second, to address the inherent biases of manual curation and to broaden coverage of potentially overlooked terms, I reviewed recent literature reporting vocabulary disproportionately employed by LLMs [12-14,18-20]. All terms meeting the original inclusion criteria from these sources were added to the initial exploratory set. The final list comprised 142 potentially AI-influenced terms (Table 1). For comparison, I created a control group of common academic words or short phrases (up to two words) in medical research by extracting items from the four-word lexical bundles identified in a previous study [21]. After excluding expressions that contained words listed as potentially AI-influenced terms in this study, such as ‘significant,’ ‘finding,’ and ‘potential,’ a final set of 84 PubMed-searchable expressions was obtained (Table 1).

**Table 1.**
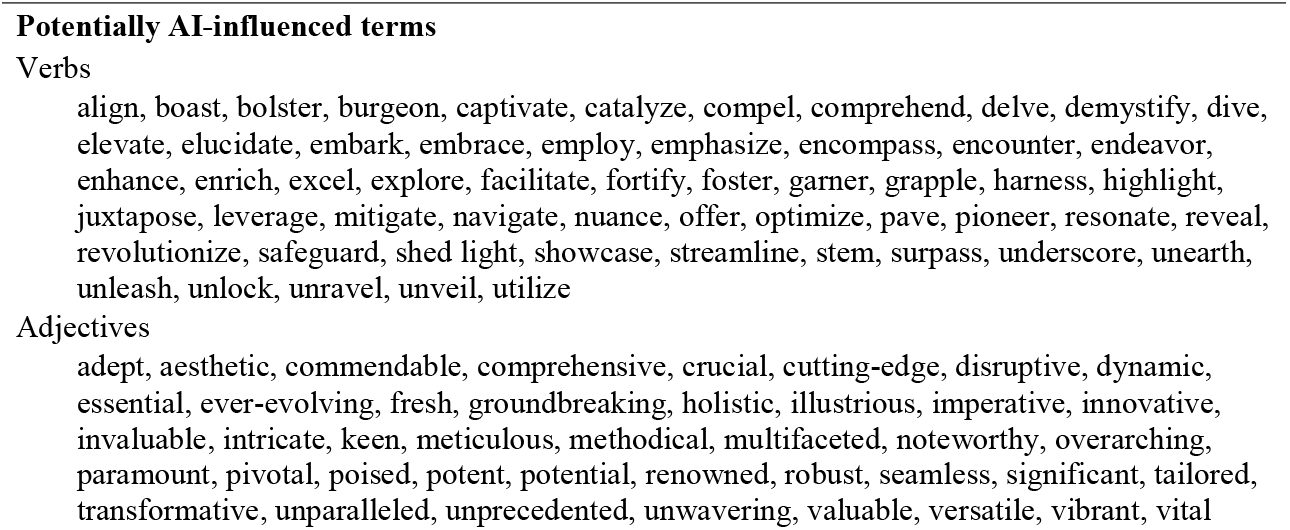
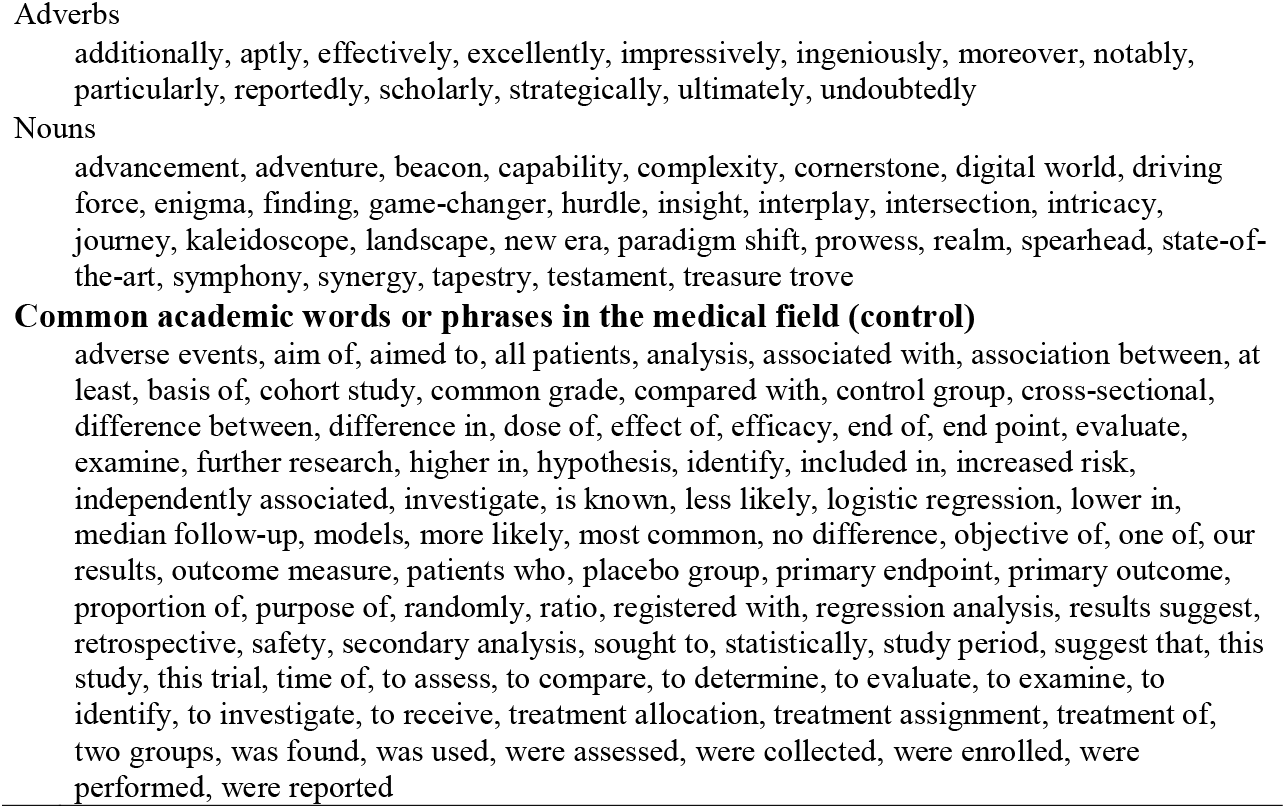
Words and phrases examined for usage rates.

I used PubMed’s advanced search feature (https://pubmed.ncbi.nlm.nih.gov/advanced/) to reveal the number of records in which these words were used by searching for “Text Word”. To ensure comprehensive coverage of verb forms in English, the search query included the base form, third person singular present, present participle/progressive, past tense, and past participle. For nouns, both singular and plural forms were incorporated. Considering the daily increase in records indexed in PubMed, the search conditions were standardized from January 1, 2000, to December 31, 2024. The search formulas for all words/phrases are shown in Appendix 1.

### Statistical Analysis

To investigate the usage trends of potentially AI-influenced terms in the PubMed database, I first calculated the usage frequency of each term by dividing the number of records containing the term by the total number of records in PubMed for each year from 2000 to 2024. This process yielded a dataset with usage frequency for each term and year.

Next, the modified Z-score transformation was used to normalize the usage frequency and facilitate comparisons across terms and years. For each term, the median and median absolute deviation (MAD) were calculated. The modified Z-score was computed by subtracting the median from each occurrence rate, dividing the result by the MAD, and multiplying by 0.6745. An absolute modified Z-score of 3.5 or higher was considered indicative of a meaningful increase or decrease in term usage to identify significant deviations [22]. Then, a linear mixed-effects model was used to compare the usage of potentially AI-influenced terms and common academic phrases from 2000 to 2024. The data, consisting of modified Z-scores for each word or phrase, were obtained and reshaped into a long format. The model, constructed using the ‘lme’ function from the ‘nlme’ package in R, included the modified Z-scores as the dependent variable, the group (potentially AI-influenced terms or common academic phrases) as a fixed effect, and a random intercept for each word or phrase to account for repeated measures. The model’s summary was generated to assess the significance of the fixed effect of the group on term usage. A line plot with 95% confidence intervals was created using the ‘ggplot2’ package to visualize the trends in mean usage for each group from 2000 to 2024. The significance level for all statistical tests was set at 0.05. These analyses were performed using R version 4.3.2.

## Results

The analysis covered a total of 27,501,542 records indexed in PubMed between January 1, 2000, and December 31, 2024. The frequency rates of each word/phrase were determined using the annual total number of records as the denominator, followed by the calculation of the modified Z-score.

In this study, among the 142 potentially AI-influenced terms verified, 100 words/phrases exhibited a modified Z-score exceeding 3.5 in 2024. The increase was particularly pronounced for many terms at the top of this list, with ‘delve,’ for instance, showing an exceptionally high Z-score. Other significantly increased terms included ‘underscore,’ ‘meticulous,’ ‘boast,’ ‘commendable,’ ‘showcase,’ ‘surpass,’ ‘intricate,’ ‘tapestry,’ and ‘emphasize’ (see Appendix 2 for the full list and Figure 1 for visual trends). While the majority of the 84 common academic phrases (controls) displayed no remarkable deviations in usage rates, ‘further research’ and ‘aim to’ also surpassed a modified Z-score of 3.5 in 2024. On the other hand, phrases such as ‘purpose of,’ ‘end of,’ ‘to determine,’ ‘hypothesis,’ ‘results suggest,’ ‘all patients,’ and ‘treatment of’ showed a notable decrease (modified Z-score < -3.5) in the same year (details in Appendix 2).

**Figure 1.**
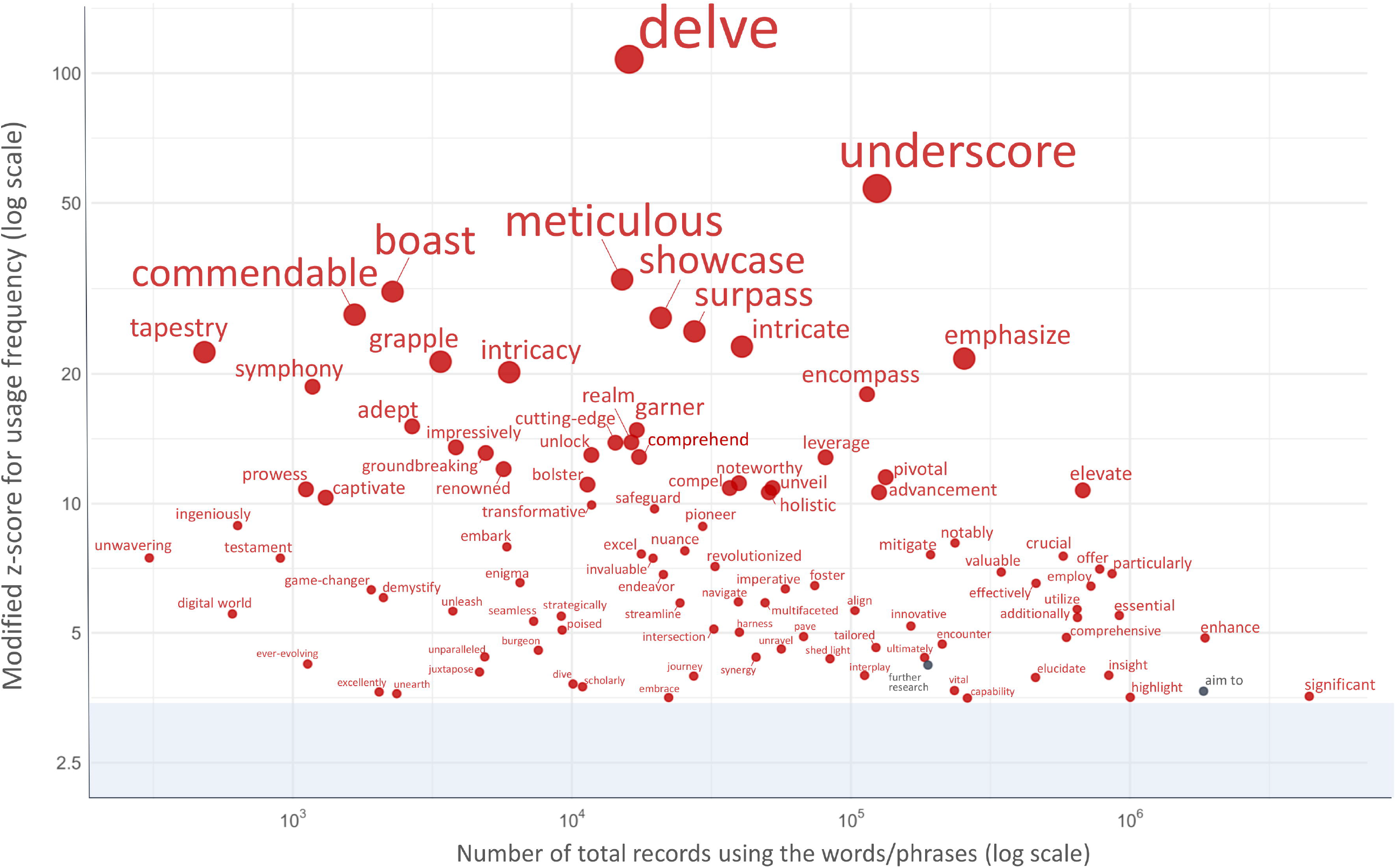
Scatter plot of word/phrase usage frequency vs. modified Z-Score in 2024. Figure 1 illustrates the relationship between the frequency of use and the modified Z-scores for words and phrases with absolute modified Z-scores exceeding 3.5 in 2024. Red circles represent potentially AI-influenced terms, while grey circles represent common academic terms in the medical field (control). The x-axis shows the number of total records using the words/phrases on a logarithmic scale, and the y-axis displays the modified Z-score for usage frequency, also on a logarithmic scale.

The linear mixed-effects model revealed a significant effect of the group (potentially AI-influenced terms vs. common academic phrases) on the modified Z-scores of term usage. The model showed that the usage of potentially AI-influenced terms was significantly higher than that of common academic phrases (β = 0.587, SE = 0.079, t(224) = 7.446, p < 0.001). The line plot (Figure 2) illustrates the trends in mean frequency for potentially AI-influenced terms and common academic phrases from 2000 to 2024. While the frequency of the control group remains relatively stable, the potentially AI-influenced terms, following a period of gradual increase, exhibit a notable and steep upward trajectory starting in 2020, which becomes particularly pronounced in 2023 and 2024.

**Figure 2.**
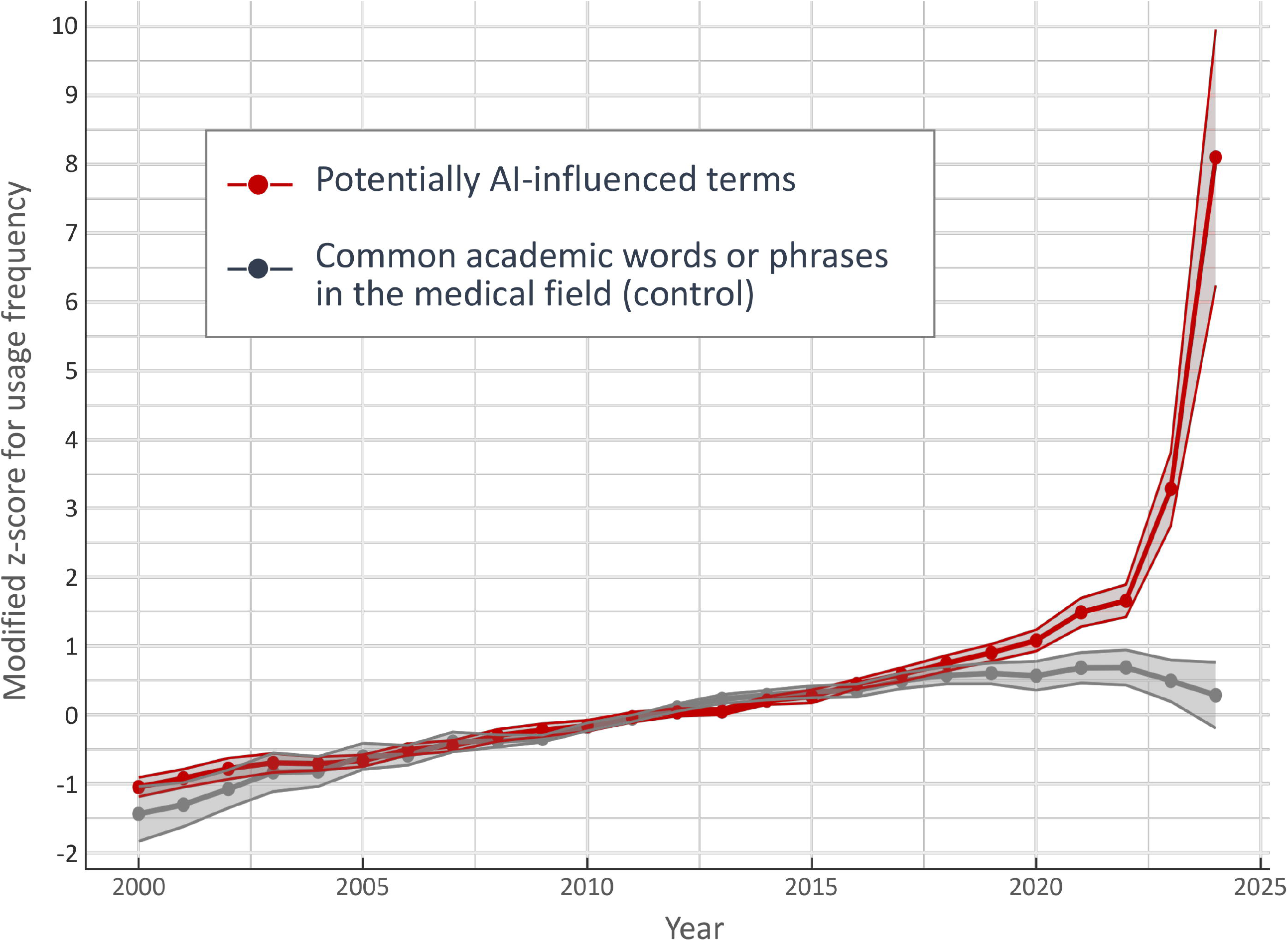
Mean usage (modified Z-scores) of potentially AI-influenced terms and common academic phrases from 2000 to 2024. Shaded areas represent 95% confidence intervals.

## Discussion

The present investigation showed that, in the fields of medicine and biology, a number of specific words and phrases, led by ‘delve,’ ‘underscore,’ ‘meticulous,’ ‘boast,’ and ‘commendable,’ have come to be used more frequently following the advent of ChatGPT. The increasing trend in the usage rates of these words/phrases was more pronounced in 2024 than in 2023 in almost all cases. This may reflect the generalization of LLM use among researchers in the fields of medicine and biology, as shown in previous findings [13]. The list of overused terms suggested in this study will help those writing with LLMs centered around ChatGPT.

It has been observed that medical texts generated by ChatGPT, while fluent and logical, tend to include less specific information and more generalized expressions compared to those authored by humans, which feature a richer and more diverse content [10]. In general papers, it has been noted that ChatGPT tends to 1) use the same style and expressions repeatedly, 2) show a decrease in the frequency of basic verbs like ‘is’ and ‘are,’ and 3) frequently use adjectives and adverbs [11]. Particularly for adjectives and adverbs, numerous words that ChatGPT frequently uses have been pointed out [14]. Because this study only counted the records where specific words or phrases occurred, it did not evaluate the weight of terms appearing multiple times. In the current study, several words that were previously identified as frequently used by ChatGPT did not exhibit a notable increase in usage; yet, ChatGPT may actually overuse these words more than suggested by the results of this study. Similarly, frequently used verbs such as ‘significant,’ ‘enhance,’ ‘essential’, and ‘particularly’ may also have been overused by LLMs more than suggested by this study.

A limitation of previous reports lies in their lack of focus on identifying specific terms whose usage has notably increased in academic writing following the widespread adoption of LLMs like ChatGPT, thus failing to comprehensively explore these characteristic terms. Specifically, the increased usage of the word ‘delve’ was incredibly pronounced compared to other words or phrases, with a modified Z-score above 100. Despite its overwhelming presence, earlier comparative analyses of human- and ChatGPT-generated texts [12-14] did not mention ‘delve,’ instead focusing on broader stylistic shifts. It is only in more recent, fine-grained corpus investigations that ‘delve’ has been flagged as a conspicuously overrepresented term in AI-assisted biomedical writing [18-20,23]; this study was among the first to identify and highlight that trend. While this study cannot conclusively establish a direct causal link between the increased use of “delve” and the emergence of ChatGPT, its influence is strongly suspected. Multiple potential factors—including training corpus composition, model architecture, tokenization methods, and context priming—could explain why ChatGPT frequently selects ‘delve.’ However, recent discussions suggested that reinforcement learning from human feedback (RLHF) might have exerted the strongest influence [20]. This indicated that human evaluation during model fine-tuning might have unintentionally encouraged stylistic word preferences, leading language models to increasingly favor particular terms such as ‘delve.’

Interestingly, some of the common academic phrases used as controls also deviated in their proportion of use in 2024. The two phrases ‘further research’ and ‘aim to’ markedly increased in frequency of use in 2024, but since they are very commonly used expressions in medical academic writing, it would be difficult for us humans to recognize that their frequency has increased. Conversely, the seven phrases ‘purpose of,’ ‘end of,’ ‘to determine,’ ‘hypothesis,’ ‘results suggest,’ ‘all patients,’ and ‘treatment of’ notably decreased in usage in 2024. When interpreting these results, we must remember that the language used in papers naturally evolves over time [24]; for example, ‘hypothesis’ and ‘results suggest’ decreased in frequency had already been declining even before the introduction of ChatGPT. In addition, the usage rate of ‘results suggest’ showed considerable variation from year to year. However, the phrase ‘all patients’ did not show a noticeable decrease in frequency of use before 2022, and its frequency of use appears to have decreased significantly after 2023 (see Appendix 2). This suggests that AI tools, such as ChatGPT, might already be subtly influencing the usage patterns of certain common academic phrases. While these phrases were selected as controls due to their frequent use in medical writing, they may have already been impacted by the emergence of AI-assisted writing.

Notably, the frequency of use for the potentially AI-influenced terms investigated in this study had already diverged markedly even before ChatGPT’s release in November 2022. The observed trends suggest a complex interplay between evolving scientific writing styles and the development of LLMs. These models, including ChatGPT, are trained on vast corpora of text data that inherently include academic and scientific writings [11,13,17]. The gradual increase in usage of certain terms prior to ChatGPT’s release may have created a recency bias in the training data, making these terms appear more relevant or current to the model. This temporal bias could have been further amplified by the RLHF during model fine-tuning [20,25]. Moreover, the increased use of specific terms in one scientific domain may have led to their adoption in other fields, a trend that LLMs could have picked up and subsequently amplified across disciplines. For instance, the word ‘delve,’ which showed a significant increase in usage in this study, seemed traditionally commonly used in the fields of computer science [26]. Furthermore, early adopters of AI writing assistants may have been exposed to and subsequently incorporated these terms into their writing, creating a human-mediated pathway for these linguistic patterns to propagate. This complex feedback loop between human writers, scientific literature, and AI language models can be more prominent in the future, specifically without being apparent to human observers. While these hypotheses remain speculative at this point, it seems highly likely that the emergence of LLMs has accelerated the naturally occurring linguistic trend changes in human scientific communication [18,19]. The subtle nature of these changes underscores the importance of continued vigilance and research to understand the evolving dynamics between AI language models and academic writing styles.

It should be noted that this study does not intend to criticize LLM use; for many researchers, especially non-native English speakers, these tools are increasingly valuable [6-8]. Rather, by identifying specific linguistic patterns—some of which, as discussed, were trending even before widespread LLM adoption—I aim to develop critical awareness. This awareness can empower medical educators and supervisors to guide authors in using LLMs more thoughtfully, refining their manuscripts to improve clarity and preserve their unique authorial voice. Educators in medical writing programs can apply these insights to encourage best practices in LLM-assisted writing; this includes guiding students and trainees to be critically aware of potentially overused terms and to actively diversify their vocabulary. Furthermore, medical educators should emphasize the importance of careful human editing to preserve the author’s unique voice and ensure originality, and promote an understanding among medical trainees that LLMs are best viewed as supportive tools for drafting and idea generation rather than as final content creators.

This study is subject to several limitations. The most important limitation is that the potentially AI-influenced terms were initially identified through a process that involved manual curation from publicly accessible online discussions. This approach inevitably introduces an element of subjectivity, particularly given the single-author nature of the study and the reliance on terms that were prominent in public discourse at the time of data collection. To address this, only terms that were independently noted across multiple unrelated sources were included, and the initial list was subsequently expanded through systematic review of recent academic literature reporting LLM-associated vocabulary. Although this methodology may have omitted certain relevant terms, the study’s aim was not to generate an exhaustive inventory of all AI-influenced expressions. Rather, it sought to test whether a representative subset of terms—frequently perceived as characteristic of LLM output—exhibited detectable shifts in real-world academic usage. The inclusion of a predefined control group was intended to contextualize these trends and help reduce the interpretive bias introduced by subjective term selection. Secondly, temporal shifts in the frequency of word or phrase use could have been influenced by external factors such as evolving research trends and shifts in the style of scientific communication, factors not accounted for in this study. The introduction of new research topics, particularly in rapidly evolving fields such as artificial intelligence, biotechnology, and pandemic-related studies, may have also contributed to these changes in language use. Thirdly, the modified Z-score was applied to the time series data. The threshold for identifying significant deviations (absolute value ≥ 3.5) was based on convention rather than strict theoretical grounds, potentially affecting the identification of significant changes. While I initially considered more complex time series models, the limited number of annual data points rendered them unsuitable, likely leading to model instability. This informed my decision to employ the modified Z-score, focusing on deviations from central tendency within each year.

## Conclusion

This study demonstrated the increased prevalence of specific words and phrases in academic writing following the introduction of ChatGPT. The list of selected terms discussed in this study can be advantageous for both users employing LLMs for writing purposes and for individuals in educational and supervisory capacities within the fields of medicine and biology. As LLMs evolve, distinguishing between human and AI-generated text may become more challenging [27]. However, simultaneously, there is a possibility that qualitative shifts in academic terminology may evolve in subtle, imperceptible ways, eluding human detection. Future research should adopt more sophisticated methods to track and understand these nuanced changes. Many researchers and medical trainees are expected to continue using LLMs for their writing—needless to say, adhering to ethical aspects and taking responsibility for the final output remain crucial aspects that medical educators must emphasize in training and mentoring when using these tools.

## Data Availability

All data and code are available at

https://github.com/matsuikentaro1/delving_into_pubmed_records.

## Data Accessibility Statement

All data and code are available at https://github.com/matsuikentaro1/delving_into_pubmed_records.

## Acknowledgements

During the preparation of this work, the author used ChatGPT (GPT-4 and GPT-4o, by OpenAI), Claude (Claude 3 Opus, Claude 3.5 Sonnet, and Claude 3.7 Sonnet, by Anthropic), and Gemini (Gemini 1.5 Pro, by Google) for specific technical assistance: drafting initial R code templates for statistical analysis, improving sentence structure for clarity, and general proofreading. These AI tools were used as writing and programming aids only, with all scientific conceptualization, interpretation of results, and conclusions being exclusively the author’s work. After using these services, the author reviewed and edited the content as needed and takes full responsibility for the content of the publication.

## Funding Information

This work was supported by the Japan Society for the Promotion of Science (JSPS) KAKENHI (grant number 22K15778).

## Ethics and Consent

Written informed consent was not applicable for this study as it involved the analysis of publicly available bibliometric data from PubMed. No individual participant data beyond publicly accessible author information was collected or analyzed, and the study did not include any potentially identifiable images or private data of individuals.

## Competing Interest

The author has no competing interests to declare.

## Notes

Source of funding support: This work was supported by the Japan Society for the Promotion of Science (JSPS) KAKENHI (grant number 22K15778).

### Competing Interest Statement

The authors have declared no competing interest.

### Funding Statement

KM is supported by the Japan Society for the Promotion of Science (JSPS) KAKENHI (grant number 22K15778).

### Summary of Updates

This version of the manuscript has been revised to update the Abstract, Discussion, and Conclusion sections to better reflect the study findings and their implications.

